# Sacral Neuromodulation in pediatric gastrointestinal motility disorders: Prospective cohort trial

**DOI:** 10.64898/2026.03.28.26349609

**Authors:** Frederike Bieling, Annemarie Kirchgatter, Angelina Bauer, Christel Weiss, Hanna Müller, Klaus E. Matzel, Andreas Rowald, Manuel Besendörfer, Sonja Diez

**Affiliations:** Friedrich-Alexander-Universität (FAU) Erlangen-Nürnberg, Department of Surgery, Paediatric Surgery, University Hospital Erlangen, Loschgestrasse 15, 91054 Erlangen, Germany; Department of Medical Statistics, Biomathematics, and Information Processing, Medical Faculty Mannheim of Heidelberg University, Theodor-Kutzer-Ufer 1-3, 68167 Mannheim, Germany; School of Biomedical Engineering and Imaging Sciences, King’s College London, Early Life Imaging Department (formerly Perinatal Imaging & Health), Block 4A, 1st Floor, South Wing, St Thomas’ Hospital SE1 7EH, London, United Kingdom; Friedrich-Alexander-Universität (FAU) Erlangen-Nürnberg, Department of Surgery, Coloproctology, University Hospital Erlangen, Maximiliansplatz 2, 91054 Erlangen, Germany; Friedrich-Alexander-Universität (FAU) Erlangen-Nürnberg, Department of Medical Informatics, Biometry and Epidemiology, Henkestraße 91, 91052 Erlangen, Germany; Department of Artificial Intelligence in Biomedical Engineering, Henkestraße 91, 91052 Erlangen, Germany

**Author notes:** **Address correspondence to:** PD Dr. med. Sonja Diez, University Hospital Erlangen, Section of Pediatric Surgery, Friedrich-Alexander-Universität (FAU) Erlangen-Nürnberg, Loschgestrasse 15, 91054 Erlangen, Germany.

**Keywords:** Sacral neuromodulation, Enteral neuromodulation, Hirschsprung, Pediatric functional constipation

## Abstract

1.

**Objectives:** To compare the efficacy and safety of invasive sacral neuromodulation (SNM) and noninvasive enteral neuromodulation (ENM) in children with refractory gastrointestinal motility disorders (GMD).

**Materials and Methods:** This prospective interventional trial enrolled pediatric patients with GMD between 2019 and 2024 at a single tertiary referral center. Children with inflammatory bowel disease or mechanical causes of GMD were excluded. Participants received either SNM via an implanted device or ENM via surface electrodes. Stimulation was delivered at 14 Hz, 210 μs pulse width, with individualized intensity (median 1.0 mA for SNM; 6.0 mA for ENM). Primary outcomes were abdominal pain, fecal incontinence, defecation frequency, and stool consistency. Treatment success was defined as clinically significant improvement in at least two of these four domains. Quality of life was assessed at baseline and 12 weeks. Safety outcomes were monitored over a 12-month follow-up.

**Results:** Of 70 eligible patients, 48 completed the study (18 SNM; 30 ENM). Diagnoses included Hirschsprung disease, functional constipation, and congenital neuronal malformations. Severe comorbidities were more frequent in the SNM group (45%) than the ENM group (3%; P = .0018). Treatment success was observed in 80% of ENM and 83% of SNM patients (P = 1.00). No significant differences were found between groups for individual outcomes. No major complications occurred. Minor adverse events were comparable (ENM 27% vs SNM 17%; P = .50).

**Conclusions:** Both SNM and ENM are effective and safe options for treating pediatric GMD and may be considered within a multimodal therapeutic approach.

## 2. Introduction

Functional gastrointestinal diseases are defined as complex bidirectional dysregulations of the brain-gut-axis in the absence of structural abnormalities.^1, 2^ During the last decade enormous progress has been made in characterizing these neuronal enteropathies not only by the connection of gastrointestinal dysmotility and psychological disturbances, but also by the complex mechanisms between inflammation and bowel barrier function, immunology and microbiome.^3^ Moreover, current research is assessing the functional neuroanatomy of the anorectum and questioning fundamental concepts of innervation and neuronal interaction in diseases.^4^ In children, these interactions might be the unifying base of gastrointestinal motility disorders, rectal outlet dysfunction and even well-known neurological disorders, e.g., Hirschsprung disease.

Current therapeutic approaches predominantly target gastrointestinal symptoms, rather than addressing this specific underlying pathophysiology.^5^ This results in high numbers of advanced stages of this primarily multidimensional disorder, including severe pain, partially irreversible rectal dilatation, decreased quality of life - comparable to organic, even oncological conditions - and psychosocial restrictions. Therapeutic failure occurs in up to one third of patients.^6^ These patients then are confronted with irreversible surgical options such as partial colectomies or ostomies.^6^ In adults, interestingly, this severely leads to potential unnecessary abdominal surgery in one third of patients.^7^

In contrast, concepts to expand this solely symptomatic treatment by the implementation of interventions targeting specific gastrointestinal control mechanisms along the brain-gut-axis are sparse. Sacral neuromodulation (SNM) is one such targeted intervention that was successfully repurposed from the field of urology to treat fecal incontinence in adult patients by aiming to recruit residual function by electrically stimulating the sacral nerves innervating pelvic floor, urinary, and anal sphincteric muscles.^8^ Today, SNM is regularly used in adult patients suffering from fecal incontinence with success; patients with constipation show mixed results.^9, 10^ In pediatric populations however, the evaluation of neuromodulation approaches in gastrointestinal motility disorders on a scientific and prospective basis is sparse.^11-16^

Concerned with the long-term implications of implanting electrodes in growing individuals, we have pioneered a non-invasive therapeutic alternative, termed enteral neuromodulation (ENM), which aims to electrically stimulate the sacral nerves with external, surface electrodes placed over the upper sacrum.^17^ Previous studies confirmed effectiveness of treatment with both ENM^11, 17^ and SNM.^18^ This study explores the comparison of these two approaches for treatment of gastrointestinal motility disorders in a prospective trial.

## 3. Patients and Methods

### A. Study design: recruitment and randomization

For this prospective, interventional trial, patients were enrolled at the pediatric surgery department at the University Hospital Erlangen (Friedrich-Alexander-Universität Erlangen-Nürnberg, Erlangen, Germany) between 2019 and 2024. The trial was designed, completed, and reported in compliance with the principles determined by the Declaration of Helsinki and the principles of Good Clinical Practice guidelines. The trial was approved by the ethic committee of the local university (No. 20_19B). It was registered at clinicaltrials.gov (ID NCT04713085).

### B. Study design: participants, data management and assessment

Eligibility was defined as follows:

- Patients aged between 4-17 years
- Symptoms compatible with the ROME IV criteria for chronic constipation ^19^ and with the ESPGHAN criteria for intractable constipation ^20^ despite optimized conservative measurements (therapy-refractory, see below)

Patients were strictly excluded from participation if mechanical causes for constipation symptoms or chronic inflammatory bowel disease were identified. Normal defecation frequency and stool consistency were defined as a frequency between 3x/week and 3x/day and a consistency of 3-4 according to the Bristol Stool Scale.^19^

After recruitment, patients and next-of-kin were not allowed to participate in any decision within the trail, but were able to withdraw participation at any timepoint. Prior to participation, next-of-kin in every case and participants > 6 years of age signed informed consent forms. The study protocol, designed according to the recommendations of the Standard Protocol Items: Recommendations for Interventionist Tests—SPIRIT^21^ and the CONSORT 2010 Statement^22^, was previously published in detail.^18^ Initial aspired randomization was not possible due to high rejection of primary surgical approach with SNM, which is reflected by the population size (ENM:SNM 2:1).

The participants’ demographics were collected in structured interviews at baseline prior to treatment, at four, eight, 12 weeks and 12 months during the therapeutic phase. Gender data were collected via self-report/report of next-of-kin. SNM was continuously applied within one year. ENM was intended to be applied for 3 months, but ENM patients were free to continue with ENM treatment after three months. Follow-up was conducted in all patients at 12 months after start of treatment. Clinical data and quality of life were assessed with specific questionnaires, bowel movement diaries, and the KINDL_R_ quality of life form.^23^ Comorbidities were hereby classified into potential low (allergies), intermediate (light psychomotor retardation) and severe influence (nonrotation, generalized neurogenic disorders) on gastrointestinal motility.

Patients and next-of-kin were finally able to choose a number between 0 (dissatisfaction with treatment) and 7 (fully satisfied with treatment) in the specific questionnaires on treatment habits.

Data assessment was based on primary outcome variables (abdominal pain, fecal incontinence, defecation frequency, and stool consistency) and secondary features (proprioception, urinary incontinency, quality of life, and safety of treatment). Patients required to monitor these outcomes on a daily interval and were classified “responding to therapy” if two out of four primary outcome variables improved >50% during the stimulation period, as specified in previous publications.^11^ Quality of life was assessed using the KINDL_R_ measurement forms.

### C. Interventions

All patients received an optimization of conservative treatment prior to participation, including non-pharmacological treatment (toilet training, lifestyle changes) and medication with oral or rectal laxatives, and were refractory to this treatment in a course of 3 months with unchanged primary outcome variables. They were advised not to change this medication during the first three months of neuromodulatory treatment. Enteral and sacral neuromodulation techniques have been described previously in detail.^11, 18^

ENM was applied via two adhesive electrodes and an external pulse generator for a transabdominal electrical field.^11, 17^ SNM was implanted in two surgical interventions: first, a tined-lead electrode with contact to sacral spinal nerve S3 or 4 was placed (see Figure 1A) and stimulated via an external pulse generator for four weeks. Afterwards, an internal pulse generator was implanted subcutaneously (see Figure 1B, both InterStim™ II/Micro SNS System (Medtronic, Minneapolis, MN).^18^

**FIGURE 1.**
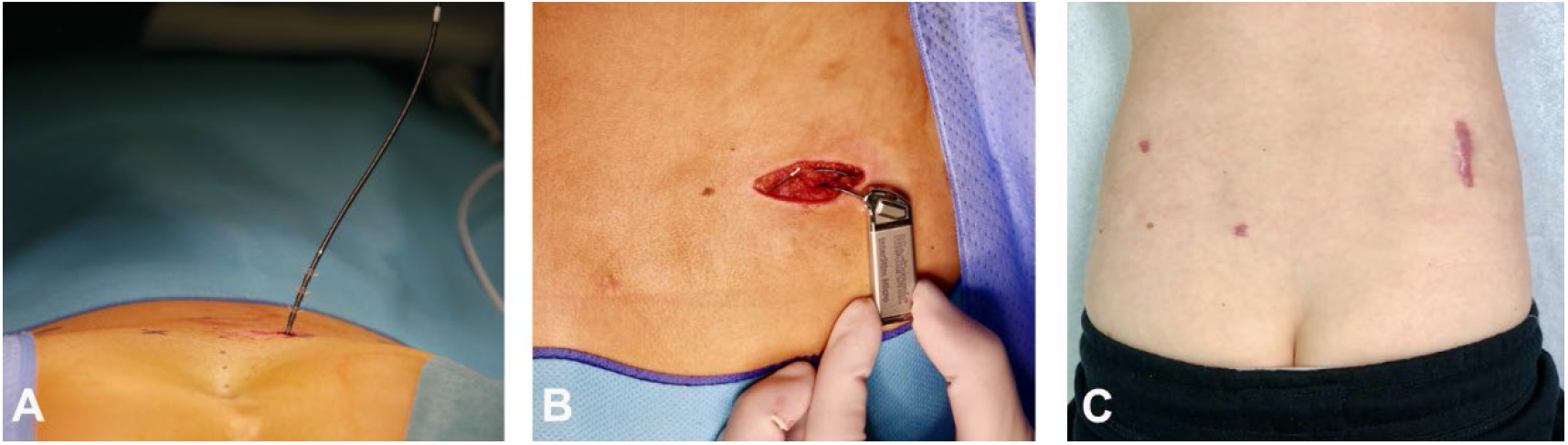
Minimally invasive SNM implantation. A. Intraoperative electrode placement, B. Intraoperative placement of the pacemaker, C. Postoperative outcome with regard to scars.

In both approaches, neurostimulation was conducted with a frequency of 14 Hz and a pulse width of 210µs. Patients were able to set stimulation intensity parameters in a pre-set range (between 0.1 and 5 (SNM)/15 (ENM) mA) based on a perianal sensation below the pain threshold and adapt during stimulation. SNM was applied continuously, while ENM could be interrupted if needed. Minimum stimulation time of ENM was 8h per day.

### D. Statistical analysis

Quantitative variables are presented as median values together with the range, categorical data are reported as frequencies and percentages. In order to compare two groups regarding a qualitative factor, Chi^2^ test or (if the preconditions of the Chi^2^ test were not fulfilled) Fisher’s exact test has been performed. For quantitative variables Mann Whitney U test has been used. For the ordinally scaled outcomes in Table 2, also medians and ranges are presented. In order to investigate changes between baseline values and values after 12 weeks, Wilcoxon tests for two paired samples have been performed. In general, the result of a statistical test has been considered as statistically significant for p values less than 0.05.

For each outcome a binary factor “Improvement” (yes / no) has been defined. In order to assess which variables may explain the improvement, multivariable logistic regression analysis have been performed, if possible. All statistical calculations have been done with SAS software (SAS Institute Inc., Cary, North Carolina, USA, release 9.4).

## 4. Results

70 patients met above defined eligibility criteria and were invited to participate in the prospective trial between Jan 01, 2019 and Dec 31, 2024. We saw an overall drop-out rate of 31% and finally completed the trail with 48 patients: SNM (n=18) or ENM (n=30). The study’s flow is illustrated in Figure 2.

**FIGURE 2.**
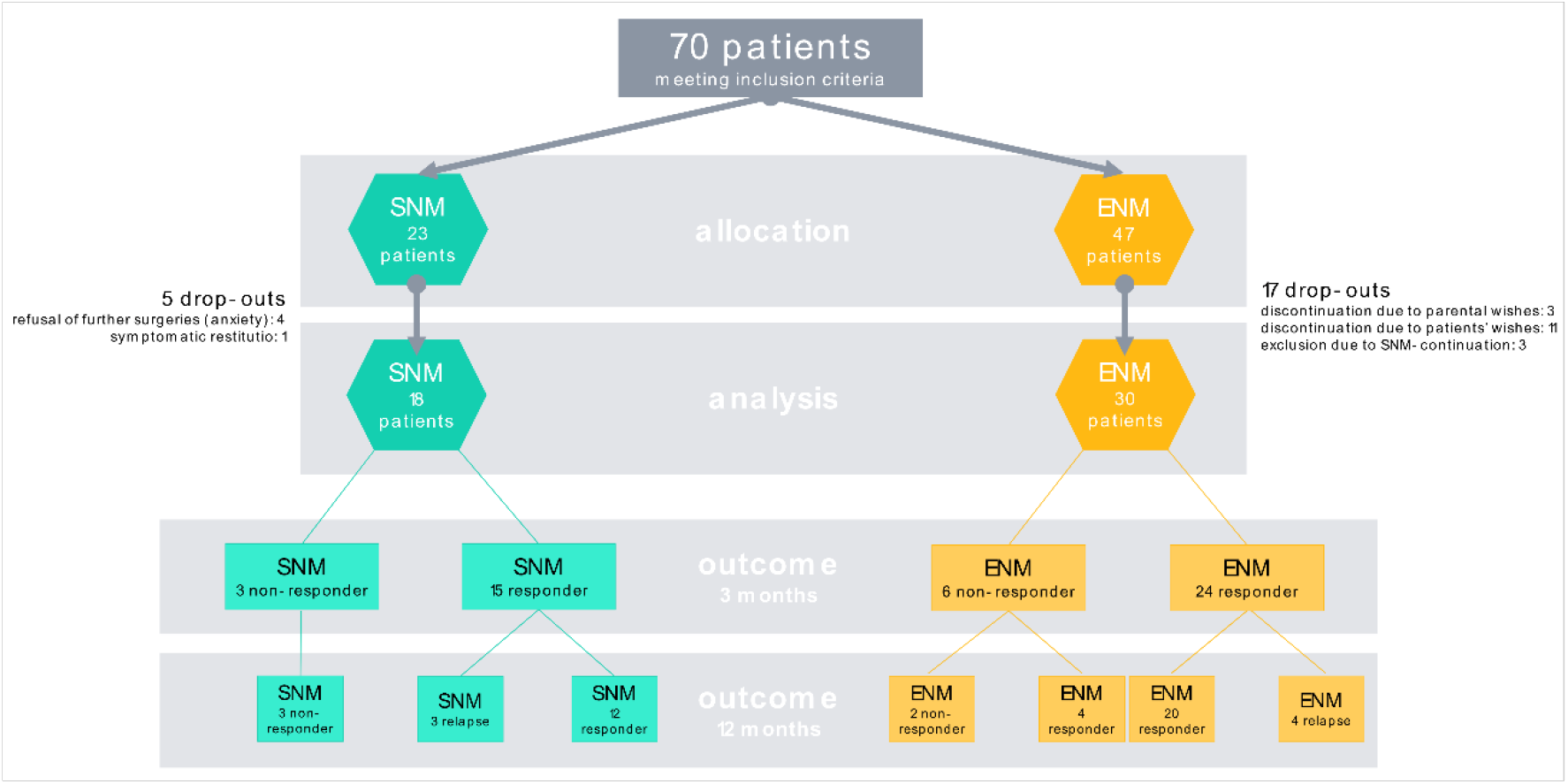
Flowchart of eligibility, inclusion, and exclusion criteria and outcome of participants. *Relapse* was defined as the loss of therapeutic response >50 % in the one-year-follow-up.

Demographics are given in Table 1. Median age of participants was 8 years (range 4-17). Age of patients and comorbidities showed significant differences between the two study groups. Patients showed deviation of normal defecation frequency and stool consistency in 100% (as defined above), abdominal pain in 60% (n=29), and fecal incontinence/soiling in 83% (n=40). SNM was implanted using the Medtronic Interstim Micro in 16/18 patients (89%). In only two patients, the Medtronic Interstim II was implanted (prior to the release of the new generation of smaller, rechargeable pulse generator). The median stimulation intensity was 1.0 mA (range 0.4 – 2.5 mA). In all patients, treatment was applied continuously without interruptions. ENM stimulation parameters were set prior to the start of the treatment for every patient. Patients were able to adjust treatment stimulation intensity and duration of application. Median stimulation intensity was 6.0 mA (range 1-15 mA). All patients applied ENM treatment within a range between 8 and 24 hours: 23/30 patients (77%) of them for >12 hours per day; seven patients (23%) between 8-12 hours per day (range 8-24 hours).

**TABLE 1.** Demographics of participants.

|  | ENM-patients<br>n=30 | SNM-patients<br>n=18 | P-value |
| --- | --- | --- | --- |
| Age [median in years (range)] | 8 (4-14) | 9.5 (5-17) | 0.0222 |
| Sex [n (%)] |  |  | 0.5271 |
| Female | 9 (30%) | 7 (39%) |  |
| Male | 21 (70%) | 11 (61%) |  |
| Diagnosis [n (%)] |  |  | 0.0125 |
| Hirschsprung disease | 9 (30%) | 9 (50%) |  |
| Functional constipation | 21 (70%) | 6 (33%) |  |
| Congenital neuronal etiologies | 0 | 3 (17%) |  |
| Comorbidities [n (%)] |  |  | 0.0014 |
| None | 19 (64%) | 4 (22%) |  |
| Low (allergy, food intolerances) | 3 (10%) | 4 (22%) |  |
| Intermediate (developmental retardation) | 7 (23%) | 2 (11%) |  |
| Severe (nonrotation, neurogenic disorders) | 1 (3%) | 8 (45%) |  |
| Perianal/rectal surgical interventions prior to study [n (%)] |  |  | 0.4682 |
| Yes | 5 (17%) | 5 (28%) |  |
| No | 25 (83%) | 13 (72%) |  |
| Medication [n (%)] |  |  | 0.8452 |
| Oral (PEG, Prucaloprid, Lactulose) | 25 (83%) | 14 (77%) |  |
| Rectal (saline/non-saline enema, Lecicarbon, anal irrigation therapy) | 2 (7%) | 1 (6%) |  |
| Combination | 3 (10%) | 3 (17%) |  |
| Fecal incontinence [n (%)] |  |  | 1.0000 |
| Yes | 25 (83%) | 15 (83%) |  |
| No | 5 (17%) | 3 (17%) |  |
| Urinary incontinence [n (%)] |  |  | 0.7624 |
| Yes | 12 (40%) | 8 (45%) |  |
| No | 18 (60%) | 10 (55%) |  |

Participants were evaluated after three months of neurostimulation, observing an overall response to therapy rate of 81% (n=39/48), meeting criteria for overall responsiveness, as defined as improvement of >50% in at least 2/4 primary outcome variables. There was no significant difference between the two study groups: ENM responders 24/30 (80%) vs. SNM responders 15/18 (83%), p=1.000. In none of the patients, a deterioration of symptoms of outcome variables did occur. Outcome variables are presented in detail in Table 2 and in Figure 3.

**FIGURE 3.**
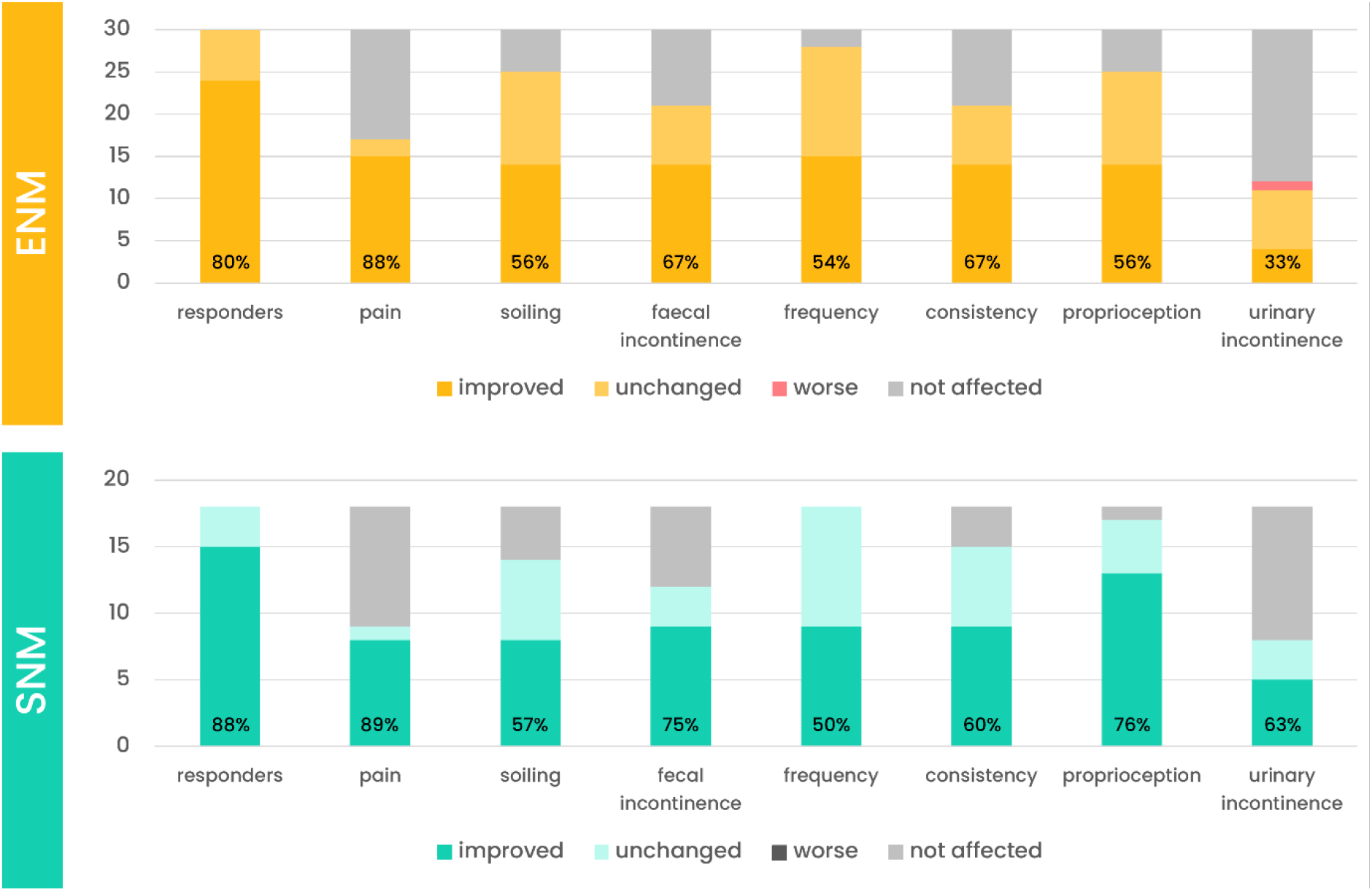
Clinical outcome variables at 12 weeks of therapy. The scale implies number of patients; percentages are given within the columns.

**TABLE 2.** The study’s outcome at three months.

|  | ENM |  |  |  | SNM |  |  |  | P-value |
| --- | --- | --- | --- | --- | --- | --- | --- | --- | --- |
|  | Number of improved patients [n (%)] | Baseline | 12 weeks | P-value ENM Baseline vs. 12 weeks | Number of improved patients [n (%)] | Baseline | 12 weeks | P-value SNM Baseline vs. 12 weeks |  |
| Abdominal pain [median (range)] | **15/17 (88%)** | 1 (0-3) | 0 (0-2) | <0.0001 | **8/9 (89%)** | 2.5 (0-3) | 1 (0-2) | 0.0195 | 1.0000 |
| Soiling [median (range)] | **14/25 (56%)** | 3 (0-3) | 2 (1-3) | 0.0554 | **8/14 (57%)** | 2.5 (1-3) | 2 (0-3) | 0.0078 | 0.9450 |
| Fecal Incontinence [median (range)] | **14/21 (67%)** | 2 (0-3) | 1 (0-3) | 0.0391 | **9/12 (75%)** | 2 (0-3) | 0 (0-3) | 0.0039 | 0.7098 |
| Frequency [median (range)] | **15/28 (54%)** | 2 (0-3) | 3 (0-3) | 0.0003 | **9/18 (50%)** | 1.5 (0-3) | 2 (1-3) | 0.0156 | 0.8219 |
| Consistency [median Bristol Stool Scale (range)] | **14/21 (67%)** | 4 (1-7) | 5 (1-7) | 0.41113 | **9/15 (60%)** | 5 (1-7) | 4 (3-7) | 0.7676 | 0.6814 |
| Proprioception [median (range)] | **14/25 (56%)** | 1 (0-3) | 1 (0-3) | <0.0001 | **13/17 (76%)** | 1 (0-3) | 2 (0-3) | 0.0002 | 0.1741 |
| Urinary Incontinence [median (range)] | **4/12 (33%)** | 2.5 (1-3) | 2 (0-3) | 0.2500 | **5/8 (63%)** | 3 (0-3) | 2 (0-3) | 0.0625 | 0.3618 |
| Quality of life [median (range)] | **6/18 (33%)** | 76.1 (59.4-96.7) | 81.3 (67.7-95.8) | 0.0718 | **4/10 (40%)** | 73.8 (48.8-96.9) | 84.4 (61.5-95.8) | 0.1094 | 1.0000 |
| Incident of safety [n (%)] | **8/30 (27%)** |  |  |  | **3/18 (17%)** |  |  |  | 0.4991 |

Comparison of all outcome variables between ENM and SNM study group. The p values in the
last column refer to the comparisons of improvements. Applied scales are classified as
abdominal pain, soiling, fecal incontinence, urinary incontinence: (0) none, (1) rare episodes
per months, (2) few episodes per week, (3) daily episodes; frequency: (0) < 2x/week, (1) 1-
2/week, (2) 1x/d, (3) 1x/d; proprioception: (0) none, (1) sporadic, (2) frequent, (3) always.
Abbreviations: ENM enteral neuromodulation (external surface electrodes). SNM sacral
neuromodulation (implanted electrodes on sacral roots).

Multiple logistic regression was conducted for all variables and revealed the neuromodulatory approach (ENM/SNM) and comorbidities as independent variables regarding improvement of proprioception with P = 0.0461 and P = 0.0472, respectively. The Odds Ratio for the neuromodulatory group (ENM/SNM) was 6.390 (CI 1.032:39.545, whereas the Odds Ratio for comorbidities was 0.493 (CI 0.245: 0.991), indicating that the higher the level of severity of comorbidities, the lower is the probability for an improvement in proprioception. The AUC of this statistical model is 0.730. Quality of life analysis did not show significances comparing baseline symptoms and 12 week-results of neuromodulation treatment (see Table 2). However, improvement of quality of life could be seen in ENM patients as a trend (P = 0.0718) and subjective treatment success was moreover assessed via questionnaires. Regarding satisfaction with treatment, ENM patients confirmed a median of 5 (range 3-7), SNM patients a median of 6 (range 4-7) with no significant differences between the two study groups (P = 0.875).

No participant showed any deterioration of primary outcome variables, constipation symptoms, pain, or neurological impairment. Negative impact of treatment was only seen in one patient under ENM therapy, who suffered from an increase of urinary incontinence. One third of ENM patients experienced skin exanthema located under the adhesive electrodes (n=8/30, 27%). This was easily resolved by exchange of electrodes to “non-allergic” variants (other adhesive components) and the slight alteration of electrode placement. In the SNM group, one patient suffered from surgical site infection, requiring removal of the infected system and implantation of a new system after three months. No further severe complications occurred in SNM patients. Minor complications included one patient with the feeling of subcutaneous tingling, almost painful sensations, which occurred only at low battery status and were resolved by improvement of charging status and another patient with electrode breakage prior to the implantation of the pulse generator, leading to re-implantation of electrode (performed simultaneously with the implantation of the pulse generator). No other surgical reinterventions were required in the patient cohort; dislocation of electrodes or dysfunctions did not occur in any participant.

Follow-up was performed in every participant within one year after baseline assessment at defined follow-up time points of four weeks, eight weeks, 12 weeks and 12 months. Improvement of primary and secondary outcome variables at 12 weeks is summarized in Table 2 and Figure 3. Compared to an overall response rate of 81% at 12 weeks, a therapeutic response of 75% at one-year follow-up could be seen (n=36/48 patients). There was no significant difference of symptom control of primary outcome variables after one year of neuromodulation treatment between the two study groups (P = 0.2300). Figure 2 summarizes the outcome of all participants. Out of 30 ENM participants, 15 (50%) were free of symptoms with or without oral laxatives after one year and further nine patients (30%) continuously confirmed improvement of symptoms >50%. However, it is notable that ENM treatment was only conducted for 12 weeks and patients were followed-up at 12 months in different conditions: 70% of ENM patients (n=21/30) were treated only conservatively at this timepoint, whereas 30% of ENM patients decided for a continuation of ENM after three months and were under neuromodulatory treatment at one-year follow-up (n=9/30). It was additionally observable that four ENM responders (n=4/24, 17%) suffered from a loss of therapeutic response at one-year follow-up, without having continued ENM therapy >12 weeks. SNM was constantly performed in every patient to the follow-up time point of one year. After one year, nine patients (50%) were symptom free and further three SNM patients (17%) confirmed improvement of symptoms >50%. The three non-responding SNM patients at 12 weeks of treatment remained non-responding at the one-year follow-up and were therefore planned for device removal. Regarding the SNM study group, three patients showed only small changes in primary outcome variables, but never reached status of “responding to treatment”. They remained non-responding to SNM treatment after one year and were therefore planned for device removal. Further three SNM patients (17%) suffered from a partial loss of therapeutic response. Even if SNM improved singular primary outcome variables or facilitated daily symptoms, all three wished for a discontinuation of SNM therapy.

## 5. Discussion

Our results demonstrate the therapeutic value of sacral neuromodulation in a pediatric patient group with gastrointestinal motility disorders within a prospective, interventional trial. Consistent with previously published data, we were able to confirm a treatment response to both, enteral neuromodulation via externally attached surface electrodes (ENM) and surgically implanted sacral neuromodulation (SNM).

Our comparative results also highlight several pragmatic aspects of this novel pediatric application. First, SNM shows lower acceptance among pediatric patients, even in the context of chronic, long-standing symptoms. The surgical approach appears to be primarily preferred for patients with comorbidities—although multiple regression analysis revealed a reduced treatment response in this subgroup regarding impaired proprioceptive function. ENM, on the other hand, demonstrates a higher susceptibility to technical failure and greater challenges in daily use. In summary, this study suggests the delineation of two distinct patient populations best suited for SNM and ENM.

Sacral neuromodulation has emerged as an uprising field of clinical and scientific research since Tanagho and colleagues established general detrusor and sphincter modulation with S3 stimulation in the 1980s.^24^ Today, SNM is considered as a promising option in the treatment of pediatric functional gastrointestinal motility disorders.^25^ Although peripheral nerve stimulation is becoming increasingly important, sacral neuromodulation accounts for only a small segment of the market, which remains largely dominated by Medtronic. Features of their newly introduced device, including a reduced size, its MRI-compatibility and the expansion of life span up to 15 years, made SNM even more appealing for the pediatric population^26^, even if off-label use is required in this age group.^27^ Although sacral neuromodulation is regarded as a promising option for pediatric motility disorders refractory to standard treatment, current evidence in children remains limited and inconsistent. Reported revision rates of up to 30% highlight the need for more robust and long-term clinical data.^25, 28^ Since 2018, our group has prospectively evaluated a non-invasive approach to replicate the effects of sacral neuromodulation in pediatric motility disorders. ENM was developed to overcome key limitations of invasive SNM—namely the need for surgery, anesthesia, and the high cost of implanted devices. Its non-invasive and low-cost nature makes ENM an attractive alternative and avoids procedure-related risks. However, whether the mechanisms of SNM translate to ENM remains unclear.

Our study is able to re-confirm the effectiveness of both, ENM and SNM treatment in therapy-refractory patients. Within the study group, we observed an excellent response of soiling and fecal incontinence up to 75% of fecal incontinence patients despite the differences to pathophysiology of adult fecal incontinence. We hypothesize that the therapeutic influence might be sufficient to influence overflow incontinence, tolerating higher rectal diameters, in combination with the improvement of defecation frequency. Defecation frequency improved in both study groups as well, resulting in a significant decrease of abdominal pain. However, urinary incontinence was only influence by SNM treatment, potentially due to the comparably more exact stimulation of the sacral spinal nerves of S3. Safety of treatment is given in both study groups, as serious complications did not occur with the exception of one surgical site infection.

The presented study contrasts the reported high proportions of surgical revisions of SNM in literature.^29^ We moreover can state that the applied neuromodulation with low frequencies does not cause severe side effects. In ENM patients, no serious adverse events occurred, patients of both groups were highly satisfied with the treatment, and the therapy was very well tolerated. However, real-life, daily application of neuromodulation in childhood certainly brings its challenges, as damage of ENM pulse generator and connections was high.

A central restriction must be acknowledged: The study was initially designed as a randomized 1:1 trial, but the allocation settings were adjusted during recruitment due to guardians’ reluctance to consent to invasive surgery once informed about the availability of a non-invasive alternative. We also observed greater acceptance of SNM among families of severely ill children, for whom a minimally invasive procedure was perceived as an acceptable burden. Our primary aim was to identify potentially meaningful differences regarding clinical outcome between the two neuromodulatory options beyond their distinct settings and procedural characteristics. Overall, comparable clinical outcomes were observed in both ENM and SNM groups. However, our findings allow for a pragmatic differentiation of treatment selection, with patient comorbidities and adaptability in daily life serving as key determinants. Importantly, similar clinical outcomes were achieved in both groups despite a potential selection bias related to comorbidity profiles, which might classify ENM as a relevant non-invasive alternative in a broader patient population.

Our study should be interpreted in light of further limitations: This is a single-center study with a limited number of patients. We included a relatively homogenous group, aiming at comparable baseline profile regarding symptoms rather than underlying pathomechanisms. These aspects may limit the generalizability of the findings to a broader population while reflecting the real-life population of specialized pediatric coloproctology centers. Another limitation must be discussed regarding the assessment and objectivation of primary outcomes. Blinding was not possible based on the study’s design and was therefore not conducted. We used validated questionnaires for most subjective variables, monitored patients rigorously for an adequate intervention and follow-up time, and assessed variables carefully based on different approaches. Objective measurements, such as pre- and post-interventional gastrointestinal transit time, colorectal distention or functional MRI, could strengthen the study’s results though.

Despite the study’s constraints, our findings contribute to the understanding of fields of application of neuromodulation in its different approaches in pediatric patient population with intractable gastrointestinal motility disorders, while proving its strengths and adding a valuable option in the pediatric clinical management. As the most recently revised German guideline on functional constipation classifies retrograde enemas and rectal manipulation as ultima ratio due to high prevalence of traumatization and offers only antegrade enemas as an alternative, which require invasive surgery for ostomies, the need for scientific evaluation of innovative approaches is obvious. ENM and SNM might be considered as therapeutic options before more invasive or potentially traumatic interventions are applied.

Emerging electrophysiological and computational research highlights fundamental mechanistic differences between invasive and non-invasive neuromodulation approaches and suggests that stimulation effects vary substantially across electrode configurations and parameter settings.^30^ Similar uncertainties apply to ENM and SNM, where mechanisms of action and therapeutic efficacy remain incompletely understood.^31^ Translating established methods from other neurostimulation modalities may help to disentangle these mechanisms and guide rational parameter selection.^32^ Considerable gaps persist: evidence on optimal stimulation parameters is limited, often confounded, and largely derived from adult cohorts; moreover, parameter tuning remains particularly challenging in pediatric patients. Key neurophysiological responses—whether directly evoked along motor axons or mediated trans-synaptically—are still poorly characterized. Future advances, including standardized electrophysiological paradigms such as paired-pulse stimulation and individualized computational modelling to refine surgical targeting, hold promise for establishing a mechanistic framework that can ultimately improve clinical outcomes in pediatric sacral neuromodulation.^33, 34^

## Conclusion

Neuromodulation represents a valuable addition to the multimodal management of pediatric gastrointestinal motility disorders. In our cohort, ENM and SNM achieved comparable clinical improvements, supporting both approaches as effective options for therapy-refractory constipation and incontinence. SNM may be preferable for children with complex comorbidities, while ENM provides a non-invasive alternative for functional constipation. Further research should identify patient characteristics that predict optimal response and refine stimulation strategies.

## Data Availability

All data produced in the present study are available upon reasonable request to the authors.

## Author statement

All authors made substantial contributions and agreed to the final version of submission.

## Clinical Trial Registration

This trial is registered at clinicaltrials.gov (ID NCT04713085, title “Sacral Neuromodulation in Children and Adolescents”). Web link: https://clinicaltrials.gov/study/NCT04713085

## Abbreviations

ENM: enteral neuromodulation (external surface electrodes)
GMD: Gastrointestinal motility disorders in children
SNM: sacral neuromodulation (implanted electrodes on sacral roots)

## Acknowledgements

This project was financially supported by the Deutsche Gesellschaft of Koloproktologie and Hannah Lerzer and Anton Rossmann, two dedicated and generous patients together with their parents, who supported the progress of our scientific work without participating in the study. Moreover, we thank all participating patients and their families for their contribution.

## 7. Appendices

**none.**

## 8. Supporting information

